# Variant connective tissue as a risk factor for Long COVID: a case-control study

**DOI:** 10.1101/2025.02.27.25323047

**Authors:** Regina A. Torok, Jeffrey Lubell, Rena M. Rudy, Jessica A Eccles, Lisa Quadt

## Abstract

**Objectives:** This study explored the extent to which two measures of joint hypermobility, a marker of variant connective tissue, predict the development of Long COVID after COVID-19 infection, and whether the severity of initial COVID-19 symptoms impacts this relationship.

**Design:** Retrospective online survey. The survey was co-designed with Long COVID patients and carers.

**Setting:** Respondents were recruited from representative online panels in the U.S. and U.K. maintained by the data firm Dynata in early 2024.

**Participants:** After data cleaning, the final dataset comprised 1,816 respondents, 352 (19.4%) who reported Long COVID and 1,464 (80.6%) who did not.

**Primary and secondary outcome measures:** The primary outcome was self-reported Long COVID, defined as experiencing symptoms related to a COVID-19 infection at least three months after the infection began. Participants were also asked to indicate whether they experienced no, mild or severe initial symptoms during each COVID-19 infection and to respond to nine questions that assessed different aspects of hypermobility: the Hakim and Grahame 5-part questionnaire (5PQ) and four additional co-produced questions. The 5PQ was used to identify individuals with generalized joint hypermobility (GJH). All nine questions were used to develop a measure of extreme hypermobility, based on the top 10^th^ percentile of hypermobility among respondents in each of six groups sorted by age and gender.

**Results:** In separate binomial logistic regressions of the dataset controlling for sex assigned at birth, age, number of infections, and number of vaccine doses, both GJH (OR 1.29, 95% CI 1.00 to 1.65) and extreme hypermobility (OR 2.12, 95% CI 1.43 to 3.16) were found to be predictive of Long COVID. Hypermobility influences the odds of getting Long COVID through two pathways. First, both GJH and extreme hypermobility increase the risk that individuals with no or moderate initial symptoms from a COVID-19 infection experience Long COVID. Second, both GJH and extreme hypermobility are significant predictors of developing severe initial symptoms from (a) COVID-19 infection(s), which is independently associated with increased Long COVID risk. A mediation analysis confirmed that extreme hypermobility influences the odds of developing Long COVID in part by increasing the likelihood that individuals experience severe initial symptoms from (a) COVID infection(s).

**Conclusions:** Both GJH and extreme hypermobility are significant risk factors for Long COVID. People with extreme hypermobility are at particularly high risk of developing Long COVID after an initial COVID-19 infection. Further research is needed to replicate these findings with other datasets and clarify the pathophysiology that explains why people with hypermobility may be at greater risk of Long COVID.

## Introduction

### Background

Long COVID (also known as “post-acute sequelae of COVID-19” (PASC), “post-acute COVID-19 syndrome” (PACS), and “post-COVID conditions” (PCC)) is a condition characterized by long-lasting persistent or newly emerging symptoms following a COVID-19 infection (see information from the World Health Organization^1^ and National Academies for Science^2^ for more background on the definition of this condition). This infection-associated chronic condition has emerged as a significant public health challenge. The most common among hundreds of documented symptoms are fatigue, post-exertional symptom exacerbation/malaise, brain fog/cognitive dysfunction, and shortness of breath.^3^ Symptoms may be new onset following initial recovery from an acute COVID-19 episode or persist from the initial illness without a gap, and they may fluctuate or exhibit a relapse/recovery pattern over time.^4^ Severity varies; some symptoms are mild, but the daily activities of around a quarter of people with Long Covid are significantly limited by their symptoms,^5^ and many people with Long Covid are completely bedbound. The burden of Long COVID is considerable, affecting an estimated 400 million people globally and costing the global economy an estimated 1 trillion USD every year.^6^ Understanding the factors that may predispose individuals to Long COVID is crucial for preventing future cases, developing effective healthcare interventions and improving the quality of life for millions affected worldwide.

Connective tissue, which is ubiquitous throughout the body, plays a critical role in maintaining the structure and function of joints, organs, muscles, bones, and nerves. Generalized joint hypermobility (GJH), which is found in approximately 20% of the UK population,^7^ is a sign of an underlying variation of connective tissue. While GJH is not always accompanied by problematic symptoms, certain clinical presentations of variant connective tissue, such as hypermobile Ehlers-Danlos Syndrome (hEDS; formerly known as EDS hypermobility type/EDS type-III) and hypermobility spectrum disorder (HSD), appear with significant clinical issues, including chronic fatigue and dysautonomia.^8^

Chronic health conditions such as fibromyalgia and myalgic encephalomyelitis/chronic fatigue syndrome (ME/CFS) frequently coexist with joint hypermobility and are often precipitated by viral infections, just as Long COVID is precipitated by SARS-CoV-2.^9–12^ ME/CFS is characterized by post-exertional malaise, disabling fatigue, unrefreshing sleep, cognitive impairment, and dysautonomia (e.g., postural orthostatic tachycardia syndrome).^13^ Dysautonomia is frequently experienced in both Long COVID and GJH,^4^ for which there is a potential mechanistic link through the effect of connective tissue on venous return.^14^ Similarities and differences between ME/CFS and Long COVID are subject to debate, but it is looking increasingly likely that a substantial subset of individuals with Long COVID have a condition that is essentially similar to or the same as ME/CFS.^15^

Genetic variants affecting collagen and extra-cellular matrix proteins, a primary component of connective tissue, may alter how the body responds to viral infections and associated inflammation. Such alterations could potentially explain why individuals with connective tissue disorders often experience chronic health conditions following viral infections. Recent case-control evidence from the British COVID Symptom Study Biobank (BCSSB) found a significant association between GJH and non-recovery from COVID-19.^16^ However, although this convenience sample was large (N=3064), it was predominantly white and female, which limits the generalizability of these results.

Given the relationships between variant connective tissue and post-viral and other chronic conditions, it is imperative to explore the connection between variant connective tissue and Long COVID in a broader sample than that described in the previous study.^16^ Furthermore, it would be of interest to determine whether there were differences related to the degree of hypermobility. This is crucial to facilitate further research and clinical intervention for this often-underserved population.

### Objectives

The primary objective of this study is to examine whether two measures of hypermobility — the established phenomenon of GJH, and our conceptualization of “extreme hypermobility” — predict the development of Long COVID. This research aims to enhance our understanding of how variant connective tissue influences the risk of Long COVID.

## Methods

### Setting

This analysis is based on data collected through a survey conducted online via the platform Qualtrics, where participants accessed study information and provided online informed consent before accessing the survey. The study was approved by the Research Ethics and Governance Committee of the Brighton and Sussex Medical School (ER/BSMS9B02/6/4).

### Public and Patient Involvement

The authorship team includes individuals with hypermobility and Long COVID, as well as a patient caregiver. The questionnaire was refined using the suggestions of additional patients with Long COVID.

### Participants

Before main recruitment, a pilot study was run on social media in December 2022 to test the feasibility of recruitment of respondents with and without Long COVID. We assessed 833 participants for eligibility, 831 of whom were determined to be eligible for the study. However, most of these participants reported having Long COVID (n=661), while recruitment of a sufficient number of respondents who did not report Long COVID (n=170) was difficult. Therefore, we chose to engage the data service firm Dynata to support recruitment from representative samples for the main study and used data from the pilot study in sensitivity analyses only. The results reported here are based on the Dynata sample alone.

Participants were drawn from online panels that are representative of the general populations of the UK and US through Dynata in January 2023. The recruitment aimed to identify at least 1,800 eligible participants, which power calculations indicated would be sufficient to detect an odds ratio of 1.19 in a logistic regression measuring the impact of GJH on Long COVID in a two-tailed test at 80% power, assuming 20% of the sample reported Long COVID and the observed frequencies of GJH in the pilot study. A total of 7,119 potential participants were screened for eligibility criteria, which included age over 18 years, fluency in English, and at least one COVID-19 infection contracted at least three months prior to taking the survey. A total of 1,816 eligible respondents completed the survey and provided responses to the key items needed for the analysis; these respondents were included in the final study sample and divided into study groups (respondents reporting Long COVID n=352, respondents not reporting Long COVID n=1465). Responses from 58 eligible respondents were excluded based on poor data quality or a non-response to a critical element needed for the analysis. Several survey questions used sliders; for these questions, a non-response was interpreted as a zero.

### Outcomes

#### Long COVID

Long COVID status was operationalized using participants’ responses to the question “Have you been diagnosed with Long COVID, or do you have symptoms that you attribute to Long COVID? For the purposes of this survey, people have Long COVID when they experience symptoms related to their COVID-19 infection at least three months after their initial infection.” Participants who selected the response “Yes, a doctor has diagnosed me with Long COVID” or “Yes, I have symptoms I attribute to Long COVID, but do not have a formal diagnosis” were identified as reporting Long COVID. Participants who selected the response “No, I do not have Long COVID” were identified as not reporting Long COVID.

#### Joint Hypermobility

Fig. 1 displays the questions and pictures used to assess joint hypermobility. GJH was assessed using the 5-part Hakim-Grahame self-report questionnaire (5PQ), where a score of 2 or greater indicates presence of GJH (Fig. 1A).^17^ Extreme hypermobility was assessed using the 5PQ and the four additional questions shown in Fig. 1B.

**Figure 1:**
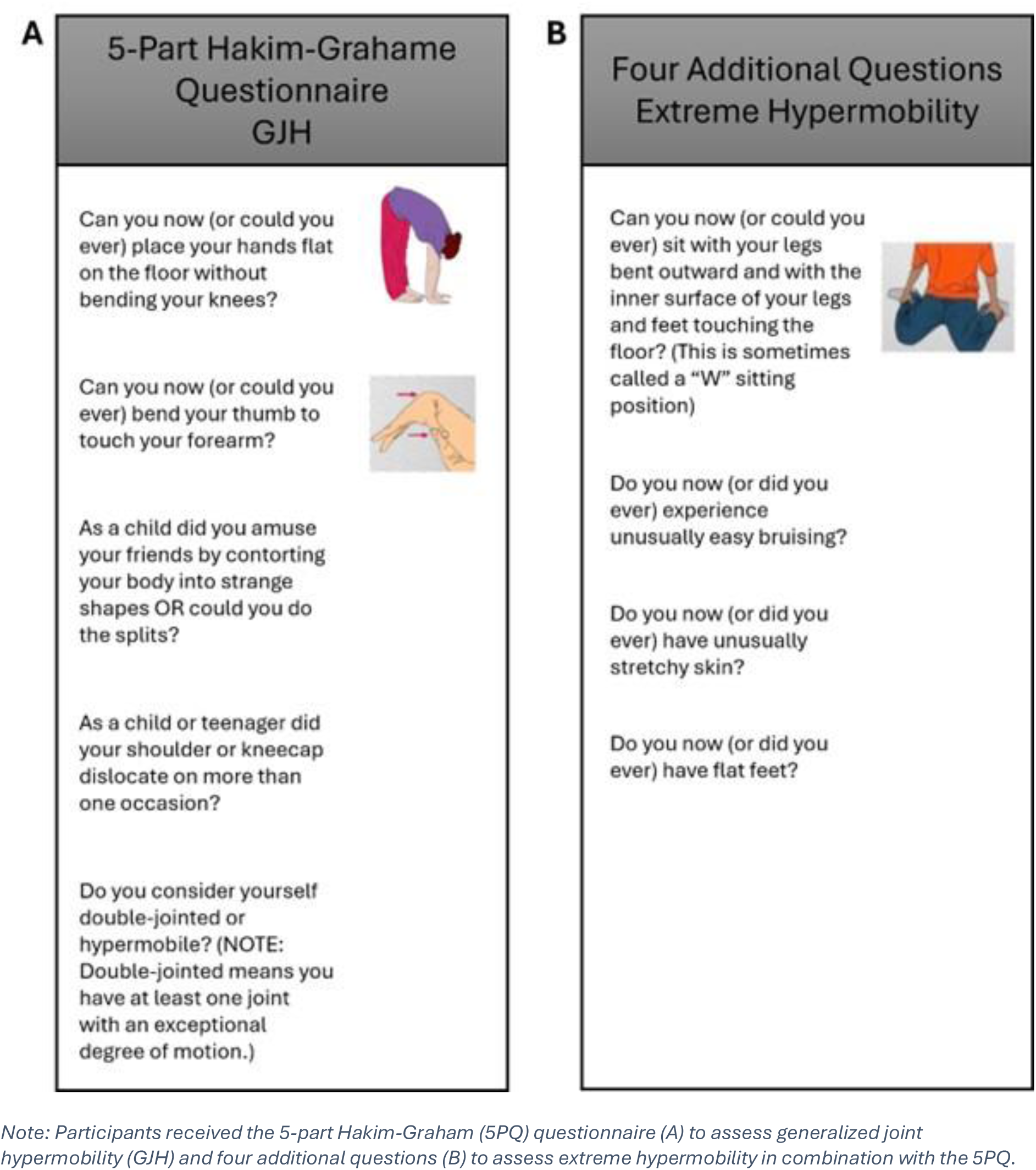
Hypermobility Assessment.

Since there is no widely accepted self-report measure of extreme hypermobility, the study team developed a novel measure based on the nine questions asked about hypermobility symptoms (Figure 1) and the core finding of a large study of 1,000 individuals that the degree of hypermobility varies with age and gender.^18^ Based on this finding, the study team stratified the sample by age (18-39, 40-59, 60+) and sex assigned at birth into six groups. The study team ordered respondents within each group according to the number of the nine hypermobility questions they answered in the affirmative and then assigned percentiles within each group reflecting each individual’s place in the distribution within that group. Those in the top 10th percentile for hypermobility within their respective age and sex group were classified as having extreme hypermobility.

### Statistical Analysis

Data analysis was performed using IBM SPSS Statistics 29, MPlus, and Jamovi version 2.3.28. Independent t-tests and Chi-square tests were used to assess basic group differences for demographic and clinical variables. The data are unweighted.

A series of separate binomial logistic regressions was used to assess whether GJH and extreme hypermobility were predictive of self-reported Long COVID. Each of these regressions controlled for age, sex at birth, number of infections (up to 3), and number of vaccine doses, factors previously shown to affect the risk of Long COVID.^19^

In an exploratory analysis, we used separate regression models for two sub-samples of respondents: (a) respondents who experienced no or mild symptoms during (all of) their COVID-19 infections(s) and (b) respondents who experienced severe symptoms (including but not limited to hospitalization) during (any of) their COVID-19 infection(s).

Binomial logistic regressions were also used to examine whether GJH and extreme hypermobility were predictive of an individual experiencing severe symptoms (including but not limited to hospitalization) during (any of) their COVID-19 infection(s). These regression models are the same as those used for the primary analyses with the exception that having severe symptoms during (any of) their COVID-19 infection(s) was used as the dependent variable rather than Long COVID.

To further test whether having severe initial symptoms mediates the relationship between extreme hypermobility and Long COVID, a mediation analysis was performed. MPlus Version 8^20^ was used to perform the mediation analysis with Long COVID (presence/absence) as the binary outcome, extreme hypermobility as the binary predictor (presence/absence), and presence or absence of severe symptomatology during any COVID-19 infection(s) as the binary mediator. Probit regression was used with full information maximum likelihood (MPlus estimator command *ML* with Monte Carlo integration), applying bootstrapping (n=2000) with the generation of 95% bias-corrected bootstrapped confidence intervals to inferentially assess direct and indirect mediation effects. Age, sex, number of infections, and number of vaccinations were included as potential confounders.

To mitigate the risk that the specific procedures used to define extreme hypermobility impacted the findings, a sensitivity analysis was conducted using an alternative definition of extreme hypermobility. In this analysis, participants who answered “yes” to 6 or more of the 9 hypermobility questions were classified as having extreme hypermobility, which identifies a similar percentage of the sample as having extreme hypermobility as the main measure.

We also repeated all analyses with a sample that combined the Dynata and the pilot study participant samples, with an added control for sample source, as a further sensitivity analysis.

## Results

### Sample Composition

After data cleaning, the sample comprised 1,816 respondents, 352 (19.4%) who reported Long COVID and 1,464 (80.6%) who did not. Demographic information is listed in Table 1. Age and sex assigned at birth differed significantly between groups and were used as covariates in further analyses. Individuals in the Long COVID group were more likely than those in the non-Long COVID group to report a parent, child or sibling with Long COVID, suggesting the possibility of a familial predisposition to the condition.

**Table 1.** Demographic Information.

| **Variable** | **Long COVID**<br>**(n=352)** | **Non-Long COVID**<br>**(n=1,464)** | **Test statistic**<br>**(df)** | **p value** |
| --- | --- | --- | --- | --- |
| Age (Mean, SD) | 49.83,<br>15.08 | 46.11, 17.15 | -4.05<br>(589.372) | **<.001** |
| Sex assigned at birth |  |  | 4.78 (1) | **.029** |
| female | 206, 58.5% | 762, 52.0% |  |  |
| male | 146, 41.5% | 702, 48.0% |  |  |
| Ethnicity |  |  | 9.21 (6) | .162 |
| Asian | 11, 3.1% | 63, 4.3% |  |  |
| Black | 25, 7.1% | 119, 8.1% |  |  |
| Hispanic/Latino | 12, 3.4% | 89, 6.1% |  |  |
| Mixed | 21, 6.0% | 60, 4.1% |  |  |
| White | 264, 75.0% | 1078, 73.6% |  |  |
| Other | 5, 1.4% | 13, 0.9% |  |  |
| Unknown | 14, 4.0% | 42, 2.9% |  |  |
| % reporting a parent, child or sibling with Long COVID | 29.6% | 11.7% | 68.32 (1) | **<.001** |

*Note: Values are n, % unless otherwise specified*

### COVID Infections

Summary data regarding the number and severity of COVID infections and number of vaccines are listed in Table 2. In line with previous findings,^21^ the Long COVID group was more likely to report two or more COVID-19 infections than the group not reporting Long COVID. We found a significant difference in symptom severity where Long COVID respondents were more likely to report experiencing severe initial symptoms during one or more of their COVID-19 infections.

**Table 2.** LC-group and non-LC-group frequencies in categories of Infections, Vaccines, initial severity, recovery, and hypermobility.

| *Variable* | **Long COVID**<br>**(n=352)** | **Non-Long COVID**<br>**(n=1,464)** | **Test statistic**<br>**(df)** | **p value** |
| --- | --- | --- | --- | --- |
| No. of COVID infections |  |  | 43.171 (2) | **<.001** |
| 1 COVID infection | 47.2% | 64.6% |  |  |
| 2 COVID infections | 36.4% | 27.4% |  |  |
| 3 or more COVID infections | 16.5% | 8.0% |  |  |
| No. of vaccine doses |  |  | 4.281 (6) | .639 |
| 2 or more vaccine doses | 73.3% | 69.7% |  |  |
| Initial symptom severity |  |  | 173.422 (4) | **<.001** |
| Mild or no initial symptoms from (all) COVID infection(s) | 39.2% | 71.7% |  |  |
| Severe initial symptoms from any COVID infection | 60.8% | 28.3% |  |  |
| Severe initial symptoms<br>(but not hospitalized) | 45.7% | 25.7% |  |  |
| Admitted to hospital but<br>not ICU | 9.1% | 2.0% |  |  |
| Admitted to ICU | 6.0% | 0.7% |  |  |
| Recovery from COVID-19 |  |  | 703.685 (5) | **<.001** |
| No or small amount of<br>recovery | 19.9% | 1.0% |  |  |
| Moderate amount of recovery | 25.3% | 1.4% |  |  |
| Substantial recovery | 21.9% | 5.1% |  |  |
| Full recovery | 32.7% | 92.1% |  |  |
| Not reporting | 0.3% | 0.3% |  |  |
| Joint hypermobility |  |  |  |  |
| GJH | 42.6% | 38.8% | 1.73 | 0.189 |
| Extreme hypermobility | 12.2% | 6.1% | 15.40 | **<0.001** |

*Note: Individual components may not add up to the total due to rounding.*

As would be expected, people reporting Long COVID more often reported they had experienced no recovery, a small recovery, or only a moderate recovery from their COVID-19 infection(s). However, about a third of those reporting having experienced Long COVID said they had fully recovered, and another 22 percent had experienced a substantial recovery by the time they completed the survey questionnaire.

### Joint Hypermobility

We found a significant group difference of presence of extreme hypermobility, but not of presence of GJH (Table 2).

### Relationship between Joint Hypermobility and Long COVID

Table 3 summarizes the results of separate serial binomial logistic regressions that assess the extent to which GJH and extreme hypermobility are predictors of Long COVID (for full results see Appendix Tables A-1 and A-2).

**Table 3:** Binomial Logistic Regressions with Long COVID as the Dependent Variable.

| **Independent Variable** | **Full Sample** |  | **Mild or No Initial Symptoms During All Infections** |  | **Severe Initial Symptoms During Any Infection** |  |
| --- | --- | --- | --- | --- | --- | --- |
|  | **Odds Ratio (95% CI)** | **P** | **Odds Ratio (95% CI)** | **P** | **Odds Ratio (95% CI)** | **P** |
| **Generalized Joint Hypermobility** | 1.29 (1.00-1.65) | .046 | 1.54 (1.05-2.25) | .027 | 0.90 (0.64-1.28)<br>(Not significant) | .571 |
| **Extreme Hypermobility** | 2.12 (1.43-3.16) | <.001 | 2.59 (1.37-4.89) | .003 | 1.33 (0.79-2.25)<br>(Not significant) | .287 |

*Note: This table reports the results of six separate binomial logistic regressions that each control for: sex at birth, age, number of infections (up to 3), and number of vaccine doses. The regressions in the first row include generalized joint hypermobility as an independent variable. The regressions in the second row include extreme hypermobility as an independent variable. The shaded portion of the graph shows the regression results for the analyses conducted to answer the primary research questions of whether GJH and extreme hypermobility are predictive of Long COVID.*

Controlling for sex at birth, age, number of infections (up to 3), and number of vaccine doses, GJH was found to be predictive of Long COVID, with an odds ratio of 1.29 (95% CI 1.00 to 1.65), and extreme hypermobility was found to be predictive of Long COVID with an odds ratio of 2.12 (95% CI 1.43 to 3.16). Age, number of infections (up to 3) and sex at birth were also significant predictors of Long COVID in both of these regressions.

The next set of binomial logistic regressions in Table 3 shows the results for individuals with mild or no initial symptoms during all infections. In this subgroup, both GJH (OR 1.54, 95% CI 1.05 to 2.25) and extreme hypermobility (OR 2.59, 95% CI 1.37 to 4.89) are significantly associated with Long COVID. Neither hypermobility measure is a significant predictor of Long COVID among the subgroup of respondents with severe initial symptoms during any COVID-19 infection.

Figure 2 illustrates an exploratory analysis of the incidence of Long COVID in the combined sample by decile of hypermobility based on the age- and gender-adjusted measure for those with mild or no initial symptoms from (all) infection(s) (blue line) and those with severe initial symptoms from any infection (orange line).

**Figure 2:**
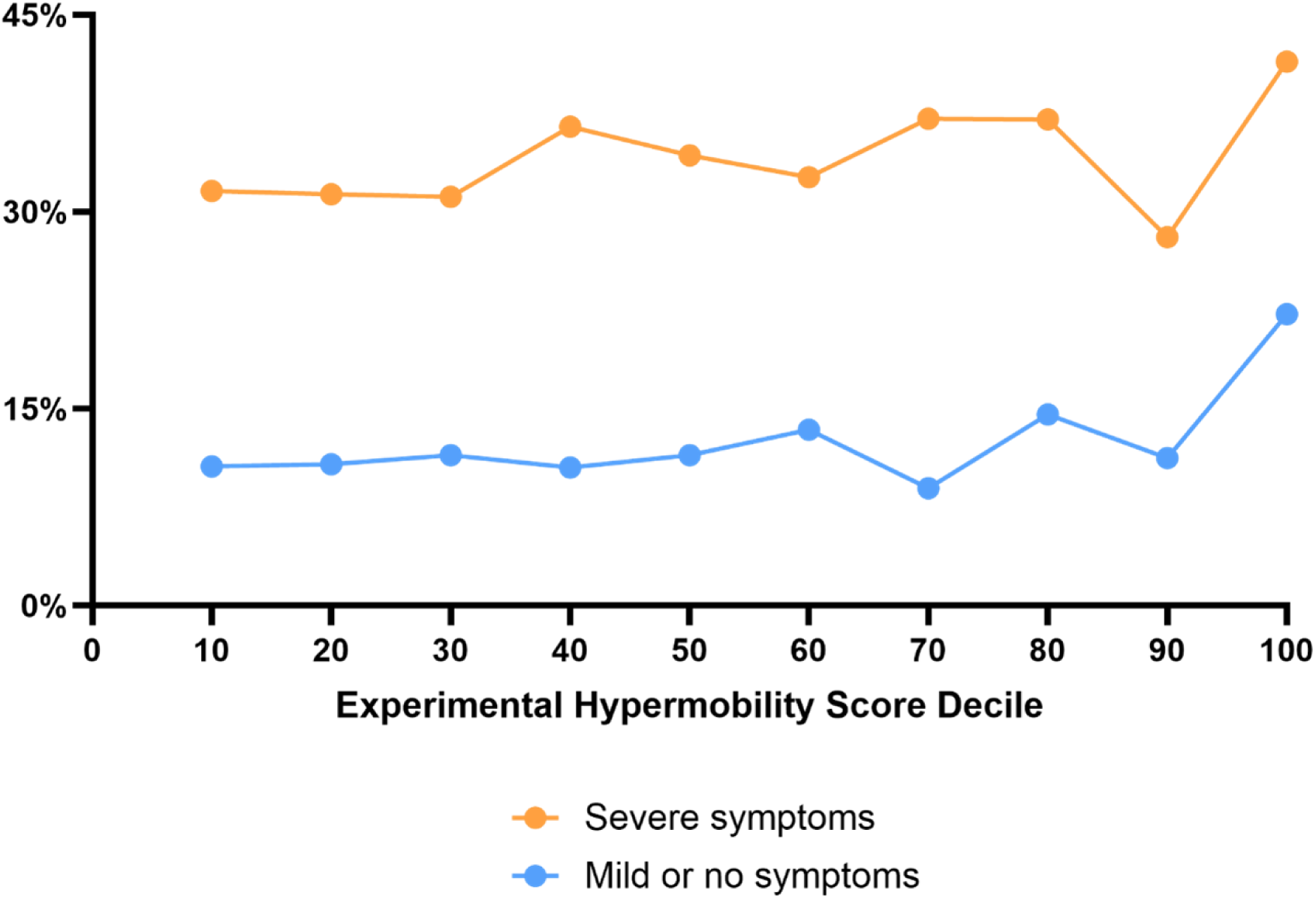
Incidence of Long COVID by (Age- and Sex-Adjusted) Decile of Hypermobility.

The exploratory analysis suggests that, in participants with no or mild initial symptoms, incidence of Long COVID appears to increase between the 30^th^ and 70^th^ percentiles, and again between the 90^th^ and 100^th^ percentiles, suggesting a ceiling effect of participants with the most extreme hypermobility. In participants with severe initial COVID-19 symptoms, we see similar trends, but with an apparent decrease in Long COVID incidence between the 80^th^ and 90^th^ hypermobility percentile. This study was not sized to detect significant differences between deciles of hypermobility; a larger study would be required to assess this issue more definitively.

### Interrelationship of Hypermobility and Severity of Initial COVID Infection(s)

GJH and extreme hypermobility appear to increase the risk of Long COVID through two mechanisms. First, as demonstrated above (Table 3), both GJH and extreme hypermobility are associated with an increased risk that people with mild or no initial symptoms from COVID-19 will develop Long COVID. Second, both GJH (OR 1.46, 95% CI 1.18 to 1.80) and extreme hypermobility (OR 2.21, 95% CI 1.53 to 3.20) are significant predictors of severe initial symptoms during one or more infection(s) in regressions controlling for sex at birth, age, number of infections (up to 3), and number of vaccine doses. Having had severe initial symptoms from any COVID-19 infection is an independent predictor of Long COVID, with an O.R. of 3.60 (95% CI 2.79 to 4.66) in a regression controlling for sex at birth, age, number of infections (up to 3), number of vaccine doses, and extreme hypermobility.

To further test whether having severe initial symptoms mediates the relationship between extreme hypermobility and Long COVID, a mediation analysis was performed. As illustrated in Figure 3, the analysis shows that extreme hypermobility is both directly associated with Long COVID and indirectly associated with Long COVID through the mediating variable of severe initial symptoms during at least one COVID-19 infection.

**Figure 3:**
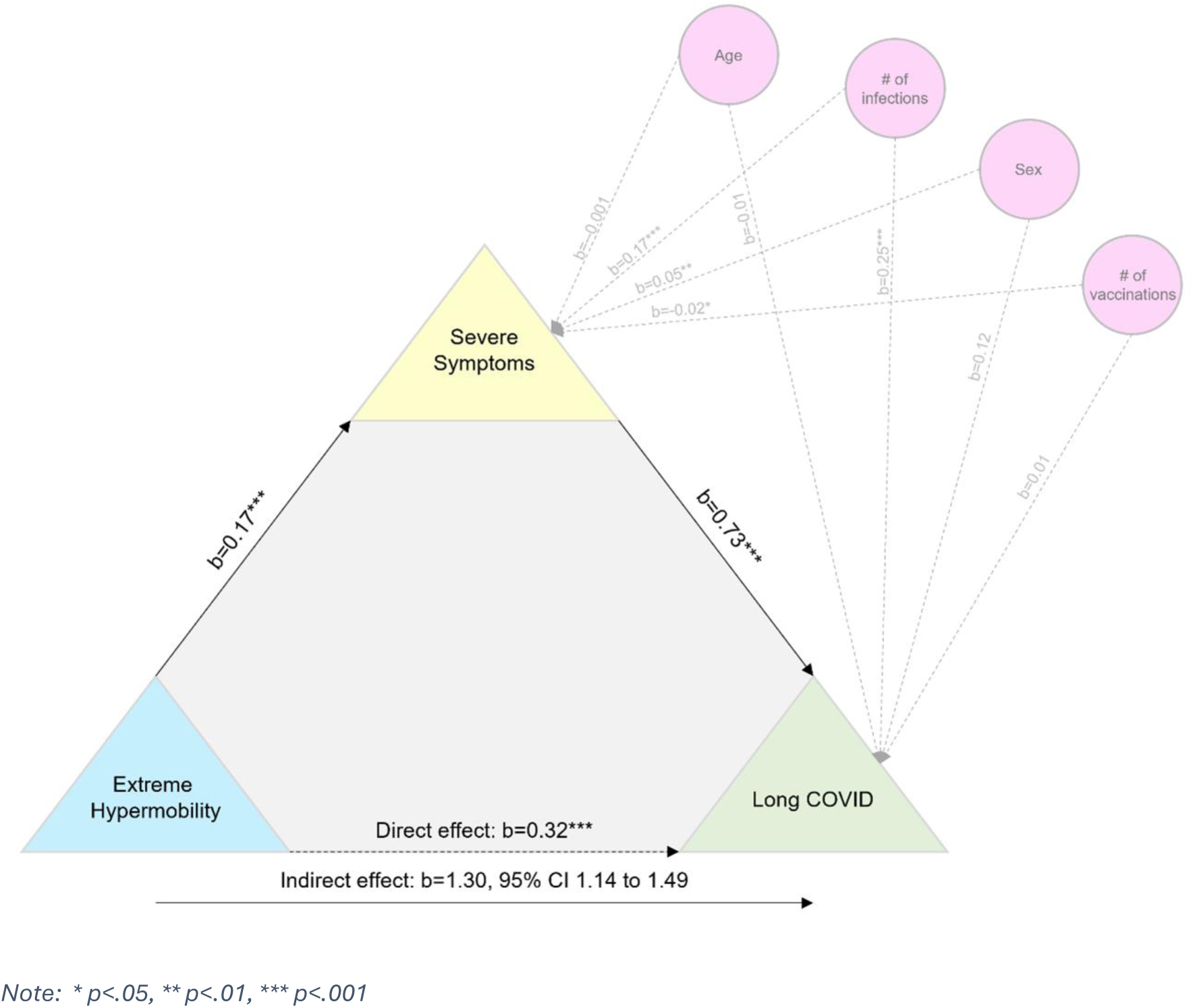
Mediation model, demonstrating mediating link of severe symptoms in the relation between extreme hypermobility and Long COVID, adjusted for potential confounders

In the mediation analysis, we found a significant relationship between number of infections and presence of Long COVID, but other covariates, including age, sex, and number of vaccine doses did not impact on development of Long COVID.

### Sensitivity Analyses

As a sensitivity analysis, the association between extreme hypermobility and Long COVID was assessed through an alternative approach that assigned everyone who answered yes to 6 or more of the 9 hypermobility questions to the category of extreme hypermobility, without sorting by age or gender. Roughly 7.4% of the combined sample meets this alternative definition of extreme hypermobility, which is very similar to the 7.3% of respondents with extreme hypermobility under this study’s primary measure. In a regression controlling for sex at birth, age, number of infections (up to 3), and number of vaccine doses, extreme hypermobility under this alternative definition was significantly associated with Long COVID with an odds ratio of 1.74 (95%, CI 1.14 to 2.65), compared with the odds ratio of 2.12 using our primary measure of extreme hypermobility.

To test the effects of omitting the pilot data collected through recruitment via social media, the binomial logistic regressions reported in this study were also performed on a dataset that combined the social media sample with the Dynata sample used in this study, with an added control for dataset. These regressions produced results that were substantially similar to those reported here. A meta-analysis is underway to analyse the association between hypermobility and Long COVID in these two samples and other relevant datasets.

## Discussion

### The Association between Hypermobility and Long COVID

The results both confirm and extend prior research. Tracking case–control evidence from the British COVID Symptom Study Biobank,^16^ this study finds GJH significantly associated with Long COVID. This study extends past research by examining the risk of Long COVID associated with having extreme hypermobility. While the 95% confidence intervals overlap, extreme hypermobility has a higher odds ratio than GJH in predicting Long COVID. Further research is needed to examine whether this difference would be statistically significant with a larger sample.

This study used a novel measure of extreme hypermobility developed specifically for this analysis. To reduce the risk that the novel definitions affected the findings, a sensitivity analysis was conducted using an alternative definition of extreme hypermobility. This sensitivity analysis produced results similar to those of the main analysis of extreme hypermobility. Further research is needed to develop a fully validated measure of extreme hypermobility and to better understand the clinical implications of this risk factor.

The results suggests that hypermobility affects the risk of Long COVID through two mechanisms. First, both measures of hypermobility are significantly associated with an increased risk of Long COVID among respondents with no or mild initial symptoms from (all) COVID-19 infection(s). Second, both measures of hypermobility are significantly associated with the likelihood of respondents experiencing severe initial symptoms from a COVID-19 infection, which is independently and strongly associated with Long COVID.

Neither of the hypermobility variables is significantly associated with respondent-reported Long COVID among the sub-group of respondents that experienced severe initial symptoms from one or more COVID-19 infections. To the extent severe initial symptoms reflect damage from a serious inflammatory cascade or other severe manifestation of COVID-19, it makes sense that people without a connective tissue disorder may be just as likely as people with a connective tissue disorder to experience long-lasting effects. This may help explain why a study of college students found no relationship between having GJH and ME/CFS following mononucleosis.^22^ Mono is essentially a severe initial reaction to infection with Epstein-Barr, and like COVID-19, it may lead to lasting effects among those with and without a connective tissue disorder. The relationship between hypermobility and Long COVID (or Mono) among those experiencing severe initial symptoms from the infection might change over time if and to the extent people with hypermobility are less likely to recover or recover more slowly from damage caused by the initial inflammatory cascade or other severe initial effect.

### Research needed to replicate results and explore relationship of connective tissue disorders and Long COVID

Further research is needed to replicate these analyses with other contexts and methods. The optimal study would have measured GJH and extreme hypermobility prospectively, before individuals are exposed to COVID-19. Since most people have been exposed to COVID-19 at least once at this point, this may be impossible to implement for COVID-19. With other pathogens on the horizon, however, prospective studies of other infection-associated chronic conditions may be possible if GJH and extreme hypermobility become more commonly measured. It will also be important to assess the risk of Long COVID and other infection-associated chronic conditions stemming from other connective tissue disorders, such as Sjogren’s syndrome, systemic lupus erythematosus, and rheumatoid arthritis.

Research is also needed to examine the extent to which individuals experience greater hypermobility after an infection and whether impacts on the extent of hypermobility are limited to or concentrated on individuals with an underlying connective tissue disorder or experienced by others. As reflected in the Supplemental Analysis, the responses of the median respondent with extreme hypermobility suggested that their current hypermobility was more or less unchanged relative to the level before their infection. However, significant shares of respondents with extreme hypermobility reported that they had grown more or less hypermobile since getting COVID-19.

Finally, research is needed to better understand why people with hypermobility may be at greater risk of developing Long COVID and what this suggests about the pathophysiology of this chronic condition. There are a number of possible explanations for why people with hypermobility may be at higher risk of Long COVID that have been suggested in or can be derived from the literature; further research is needed to determine which, if any, are valid. Some have suggested that vascular laxity makes people with Ehlers-Danlos Syndrome vulnerable to Postural Orthostatic Tachycardia Syndrome (POTS) due to the distension of veins from hydrostatic pressure, which increases venous pooling,^23^ ^24^ which is characteristic of a ME/CFS-type presentation of Long COVID. An alternative explanation is that the interrelationship of connective tissue and mast cells leads people with Ehlers-Danlos Syndrome or hypermobility spectrum disorder to be particularly vulnerable to mast cell activation disorders,^25^ which can in turn prompt both POTS and conditions like Long COVID and ME/CFS.^26^ ^27^ A third possibility is that the increased risk of Long COVID is related to the susceptibility of people with Ehlers-Danlos syndrome to autoimmune disorders;^28^ an increasing body of evidence documents the role that autoantibodies play in Long COVID.^29^ ^30^

A fourth possibility is that people with connective tissue disorders are more likely to experience, and slower to recover from, the vascular damage that characterizes Long COVID and associated damage to the extracellular matrix.^31^ ^32^ Fifth, the increased Long COVID risk from hypermobility could be related to the tendency of people with Ehlers-Danlos syndrome to experience spinal problems such as Chiari malformation and craniocervical instability.^10^ ^33^ ^34^ As vascular and blood-brain-barrier permeability increases among people with Long COVID due to inflammation and vascular damage,^35^ ^36^ the resulting leakage of protein-rich fluid in the brain can increase the volume of glymphatic and cerebrospinal fluid in need of drainage. Spinal problems such as Chiari malformation and craniocervical instability can impair the drainage of cerebrospinal fluid, resulting in an increase in intracranial pressure that leads to brainstem compression;^37^ an impaired glymphatic system may compound the problem.^31^ ^38^ Drawing on the experience of patients in their practice, Maxwell and Wardly have proposed an overarching hypothesis that ties together several of these mechanisms, including mast cell activation, hypermobility, dysautonomia and spinal complications.^39^

These and other possibilities should be rigorously assessed to determine whether they are valid explanations for the increased risk of Long COVID associated with hypermobility. This research may shed important light on the field’s understanding of the pathophysiology of Long COVID, which remains incomplete.

### Strengths and Limitations

This study has several strengths. First, recruitment from representative online panels means that this study’s results are more generalizable to the adult population than studies with convenience samples. Second, employing a validated diagnostic questionnaire for hypermobility (the 5PQ) ensures that this study is comparable with others that use this measure. Third, the additional questions used to operationalize extreme hypermobility have the advantage of addressing hypermobility in body parts (feet and skin) and orientations (W-sitting) not considered in the 5PQ. The concept of extreme hypermobility has not been previously studied and represents an important innovation of this study; we found our proposed measure of extreme hypermobility to be a better predictor of Long COVID morbidity than the more standard GJH. A key limitation of this study is the reliance on self-reported data collected at a single point in time, rather than longitudinally. Respondents may or may not have a good recollection of their condition before getting COVID-19. In addition, the measure of extreme hypermobility is novel and not yet validated.

### Conclusion

These findings suggest that people with variant connective tissue (manifest as joint hypermobility) disproportionately experience Long COVID. Long COVID among people with variant connective tissue may potentially constitute a sub-type of Long COVID, a heterogenous condition. Better recognition and enhanced understanding of this association is necessary to address the public health challenges of Long COVID and enhance personalised care of often underserved groups.^40^

## Data Availability

Data will be made available upon reasonable request to the corresponding author.

## Funding

This study was partially funded by the Ehlers Danlos Syndrome Research Foundation and The Ehlers Danlos Society. However, the funders played no role in the research, which was conducted independently by the authors.

## Acknowledgments

We are grateful to Eric Hedberg for valuable statistical advice and review of the manuscript and to the individuals with Long COVID who provided feedback on the initial questionnaire and shared the pilot questionnaire via Social Media.

## Author Contributions

All authors conceived, planned, and designed this study. JL, JAE, and LQ obtained funding. RT and LQ collected data. RAT and JL conducted data analyses. RAT, JL, JAE, and LQ interpreted data. RAT and JL drafted the full manuscript. JAE, LQ, and RR edited the manuscript. All authors approved the final version.

The lead authors attest that all listed authors meet authorship criteria and that no others meeting the criteria have been omitted.

## Competing interests

All authors have completed the **Unified Competing Interest form** (available on request from the corresponding author) and declare: funding for the research noted above in the Funding Section; no financial relationships with any organisations that might have an interest in the submitted work in the previous three years, and no other relationships or activities that could appear to have influenced the submitted work.

## Ethics Approval

The study was approved by the Research Ethics and Governance Committee of the Brighton and Sussex Medical School (ER/BSMS9B02/6/4).

## Transparency Statement

The lead authors affirm that the manuscript is an honest, accurate, and transparent account of the study being reported; that no important aspects of the study have been omitted; and that any discrepancies from the study as planned (and, if relevant, registered) have been explained.

## Data sharing statement

Data will be made available upon reasonable request to the corresponding author.

## Appendix Tables

**Table A-1:** Binomial Logistic Regression exploring association between GJH and reporting Long COVID for the full sample.

| Predictor | Estimate | SE | Z | p | Odds ratio | 95% Confidence Interval |  |
| --- | --- | --- | --- | --- | --- | --- | --- |
|  |  |  |  |  |  | Lower | Upper |
| Intercept | -3.6251 | 0.29257 | 12.391 | < .001 | 0.0266 | 0.0150 | 0.0473 |
| GJH | 0.2537 | 0.12707 | 1.996 | 0.046 | 1.2887 | 1.0046 | 1.6532 |
| Sex at birth | 0.2436 | 0.12328 | 1.976 | 0.048 | 1.2758 | 1.0020 | 1.6245 |
| Age | 0.0208 | 0.00408 | 5.086 | < .001 | 1.0210 | 1.0128 | 1.0292 |
| Number of COVID-19 infections (up to 3) | 0.6350 | 0.08686 | 7.310 | < .001 | 1.8870 | 1.5916 | 2.2372 |
| Number of vaccine doses | -0.0103 | 0.03999 | -0.259 | 0.796 | 0.9897 | 0.9151 | 1.0704 |

**Table A-2.** Binomial Logistic Regression exploring association between extreme hypermobility and reporting Long COVID for the full sample.

| Predictor | Estimate | SE | Z | p | Odds ratio | 95% Confidence Interval |  |
| --- | --- | --- | --- | --- | --- | --- | --- |
|  |  |  |  |  |  | Lower | Upper |
| Intercept | -3.5168 | 0.27852 | -12.627 | < .001 | 0.0297 | 0.0172 | 0.0513 |
| Extreme hypermobility | 0.7537 | 0.20211 | 3.729 | < .001 | 2.1248 | 1.4298 | 3.1576 |
| Sex at birth | 0.2583 | 0.12334 | 2.094 | 0.036 | 1.2947 | 1.0167 | 1.6488 |
| Age | 0.0195 | 0.00400 | 4.865 | < .001 | 1.0196 | 1.0117 | 1.0277 |
| Number of COVID-19 Infections (up to 3) | 0.6292 | 0.08716 | 7.218 | < .001 | 1.8760 | 1.5814 | 2.2255 |
| Number of vaccine doses | -0.0137 | 0.04003 | -0.343 | 0.732 | 0.9864 | 0.9119 | 1.0669 |

## Notes

### Competing Interest Statement

The authors have declared no competing interest.

### Author Declarations

The Research Ethics and Governance Committee of the Brighton and Sussex Medical School gave ethical approval for this work (ER/BSMS9B02/6/4).

## References

1. World Health Organization. A clinical case definition of post COVID-19 condition by a Delphi consensus. https://iris.who.int/bitstream/handle/10665/345824/WHO-2019-nCoV-Post-COVID-19-condition-Clinical-case-definition-2021.1-eng.pdf?sequence=12021.

2. National Academies of Sciences E, and Medicine. A Long COVID Definition: A Chronic, Systemic Disease State with Profound Consequences. Washington, DC: The National Academies Press 2024.

3. Davis HE, Assaf GS, Mccorkell L, et al. Characterizing long COVID in an international cohort: 7 months of symptoms and their impact. eClinicalMedicine 2021;38:101019. doi: 10.1016/j.eclinm.2021.101019

4. Davis HE, Mccorkell L, Vogel JM, et al. Long COVID: major findings, mechanisms and recommendations. Nature Reviews Microbiology 2023 doi: 10.1038/s41579-022-00846-2

5. Ford ND, Slaughter D, Edwards D, et al. Long COVID and Significant Activity Limitation Among Adults, by Age — United States, June 1–13, 2022, to June 7–19, 2023. MMWR Morbidity and Mortality Weekly Report 2023;72(32):866–70. doi: 10.15585/mmwr.mm7232a3

6. Al-Aly Z, Davis H, Mccorkell L, et al. Long COVID science, research and policy. Nature Medicine 2024;30(8):2148–64. doi: 10.1038/s41591-024-03173-6

7. Mulvey MR, Macfarlane GJ, Beasley M, et al. Modest association of joint hypermobility with disabling and limiting musculoskeletal pain: results from a large-scale general population-based survey. Arthritis care & research 2013;65(8):1325–33. doi: 10.1002/acr.21979 [published Online First: 2013/02/13]

8. Castori M, Tinkle B, Levy H, et al. A framework for the classification of joint hypermobility and related conditions. Am J Med Genet C Semin Med Genet 2017;175(1):148–57. doi: 10.1002/ajmg.c.31539

9. Eccles JA, Thompson B, Themelis K, et al. Beyond bones: The relevance of variants of connective tissue (hypermobility) to fibromyalgia, ME/CFS and controversies surrounding diagnostic classification: an observational study. Clinical Medicine 2021;21(1):53–58. doi: 10.7861/clinmed.2020-0743

10. Bragée B, Michos A, Drum B, et al. Signs of Intracranial Hypertension, Hypermobility, and Craniocervical Obstructions in Patients With Myalgic Encephalomyelitis/Chronic Fatigue Syndrome. Frontiers in Neurology 2020;11 doi: 10.3389/fneur.2020.00828

11. Castori M, Celletti C, Camerota F, et al. Chronic fatigue syndrome is commonly diagnosed in patients with Ehlers-Danlos syndrome hypermobility type/joint hypermobility syndrome. Clin Exp Rheumatol 2011;29(3):597–8. [published Online First: 2011/07/05]

12. Renz-Polster H, Tremblay M-E, Bienzle D, et al. The Pathobiology of Myalgic Encephalomyelitis/Chronic Fatigue Syndrome: The Case for Neuroglial Failure. Frontiers in Cellular Neuroscience 2022;16 doi: 10.3389/fncel.2022.888232

13. Committee on the Diagnostic Criteria for Myalgic Encephalomyelitis/Chronic Fatigue S, Board on the Health of Select P, Institute of M. The National Academies Collection: Reports funded by National Institutes of Health. Beyond Myalgic Encephalomyelitis/Chronic Fatigue Syndrome: Redefining an Illness. Washington (DC): National Academies Press (US) Copyright 2015 by the National Academy of Sciences. All rights reserved. 2015.

14. Midtlien JP, Curry BP, Chang E, et al. Characterizing a new clinical phenotype: the co-existence of cerebral venous outflow and connective tissue disorders. Frontiers in Neurology 2024;14 doi: 10.3389/fneur.2023.1305972

15. Weigel B, Eaton-Fitch N, Thapaliya K, et al. Illness presentation and quality of life in myalgic encephalomyelitis/chronic fatigue syndrome and post COVID-19 condition: a pilot Australian cross-sectional study. Quality of Life Research 2024 doi: 10.1007/s11136-024-03710-3

16. Eccles JA, Cadar D, Quadt L, et al. Is joint hypermobility linked to self-reported non-recovery from COVID-19? Case–control evidence from the British COVID Symptom Study Biobank. BMJ Public Health 2024;2(1):e000478. doi: 10.1136/bmjph-2023-000478

17. Hakim A, Grahame R. A SIMPLE QUESTIONNAIRE TO DETECT HYPERMOBILITY: AN ADJUNCT TO THE ASSESSMENT OF PATIENTS WITH DIFFUSE MUSCULOSKELETAL PAIN. International Journal of Clinical Practice 2003;57(3):163–66. doi: 10.1111/j.1742-1241.2003.tb10455.x

18. Singh H, McKay M, Baldwin J, et al. Beighton scores and cut-offs across the lifespan: cross-sectional study of an Australian population. Rheumatology (Oxford) 2017 doi: 10.1093/rheumatology/kex043

19. Tsampasian V, Elghazaly H, Chattopadhyay R, et al. Risk Factors Associated With Post−COVID-19 Condition. JAMA Internal Medicine 2023;183(6):566. doi: 10.1001/jamainternmed.2023.0750

20. Muthén L, Muthén B. Statistical Analysis with Latent Variables: Mplus User’s Guide: Muthén & Muthén, Los Angeles, CA, 2012.

21. Bowe B, Xie Y, Al-Aly Z. Acute and postacute sequelae associated with SARS-CoV-2 reinfection. Nature Medicine 2022;28(11):2398–405. doi: 10.1038/s41591-022-02051-3

22. Poomkudy JT, Torres C, Jason LA, et al. Joint Flexibility and Myalgic Encephalomyelitis/Chronic Fatigue <u>Syndrome</u> After Mononucleosis. Clin Ther 2024 doi: 10.1016/j.clinthera.2023.12.011 [published Online First: 20240118]

23. Rowe PC, Barron DF, Calkins H, et al. Orthostatic intolerance and chronic fatigue syndrome associated with Ehlers-Danlos syndrome. The Journal of pediatrics: Mosby Inc., 1999:494–99.

24. Miller AJ, Stiles LE, Sheehan T, et al. Prevalence of hypermobile Ehlers-Danlos syndrome in postural orthostatic tachycardia syndrome. Autonomic Neuroscience: Basic and Clinical 2020;224 doi: 10.1016/j.autneu.2020.102637

25. Monaco A, Choi D, Uzun S, et al. Association of mast-cell-related conditions with hypermobile syndromes: a review of the literature. Immunologic Research 2022;70(4):419–31. doi: 10.1007/s12026-022-09280-1

26. Batiha GE-S, Al-Kuraishy HM, Al-Gareeb AI, et al. Pathophysiology of Post-COVID syndromes: a new perspective. Virology Journal 2022;19(1) doi: 10.1186/s12985-022-01891-2

27. Jahanbani F, Sing JC, Maynard RD, et al. Longitudinal cytokine and multi-modal health data of an extremely severe ME/CFS patient with HSD reveals insights into immunopathology, and disease severity. Frontiers in Immunology 2024;15 doi: 10.3389/fimmu.2024.1369295

28. Rodgers KR, Gui J, Dinulos MBP, et al. Ehlers-Danlos syndrome hypermobility type is associated with rheumatic diseases. Scientific Reports 2017;7(1):39636. doi: 10.1038/srep39636

29. Chen H-J, Appelman B, Willemen H, et al. Transfer of IgG from Long COVID patients induces symptomology in mice: Cold Spring Harbor Laboratory, 2024.

30. Santos Guedes de Sa K, Silva J, Bayarri-Olmos R, et al. A causal link between autoantibodies and neurological symptoms in long COVID. medRxiv 2024:2024.06.18.24309100. doi: 10.1101/2024.06.18.24309100

31. Wu L, Zhang Z, Liang X, et al. Glymphatic system dysfunction in recovered patients with mild COVID-19: A DTI-ALPS study. iScience 2024;27(1):108647. doi: 10.1016/j.isci.2023.108647

32. Lubell J. People with a connective tissue disorder may be especially vulnerable to the endothelial damage that characterizes long COVID due to the fragility of their vasculature and slow wound healing. Angiogenesis 2024 doi: 10.1007/s10456-024-09908-w

33. Friedlander RM. Congenital and Acquired Chiari Syndrome. New England Journal of Medicine 2024;390(23):2191–98. doi: 10.1056/NEJMra2308055

34. Milhorat TH, Bolognese PA, Nishikawa M, et al. Syndrome of occipitoatlantoaxial hypermobility, cranial settling, and Chiari malformation Type I in patients with hereditary disorders of connective tissue. Journal of Neurosurgery: Spine SPI 2007;7(6):601–09. doi: 10.3171/SPI-07/12/601

35. Greene C, Connolly R, Brennan D, et al. Blood–brain barrier disruption and sustained systemic inflammation in individuals with long COVID-associated cognitive impairment. Nature neuroscience 2024;27(3):421–32. doi: 10.1038/s41593-024-01576-9

36. VanElzakker MB, Bues HF, Brusaferri L, et al. Neuroinflammation in post-acute sequelae of COVID-19 (PASC) as assessed by [11C]PBR28 PET correlates with vascular disease measures. Brain, Behavior, and Immunity 2024;119:713–23. doi: 10.1016/j.bbi.2024.04.015

37. Henderson FC, Schubart JR, Narayanan MV, et al. Craniocervical instability in patients with Ehlers-Danlos syndromes: outcomes analysis following occipito-cervical fusion. Neurosurgical Review 2024;47(1) doi: 10.1007/s10143-023-02249-0

38. Zhou Z, Zhao T, Sekendiz Z, et al. Impaired Cerebrospinal Fluid 18F-FEPPA Clearance in Long COVID Suggests Altered Glymphatic System Function. Journal of Nuclear Medicine 2024;65(supplement 2):242407.

39. Maxwell AJ. The Complex Path to Intracranial Hypertension and CSF Leak in those with Hypermobility and Dysautonomia; The Theory of Spiky-Leaky Syndrome. MAR Pediatrics 2024;5(3)

40. Halverson CME, Penwell HL, Francomano CA. Clinician-associated traumatization from difficult medical encounters: Results from a qualitative interview study on the Ehlers-Danlos Syndromes. SSM - Qualitative Research in Health 2023;3:100237. doi: 10.1016/j.ssmqr.2023.100237

