## Supplemental Table 1 for "Variant connective tissue as a risk factor for Long COVID: a case-control study"

**Supplemental Analysis**

**Changes in hypermobility symptoms since getting COVID-19**

Supplemental Table 1 shows changes in respondents’ ratings (from 0-100) of four hypermobility signs for two time periods (a) before getting COVID-19 and (b) as of the time of the survey. The four symptoms are: bendiness/double-jointedness, "clicky" or dislocating joints, bruising easily, and having unusually stretch skin. Among Long COVID respondents, the median response shows little or no change from over this time period among both those with extreme hypermobility and those without it. In examining the distribution of change over time, the group with extreme hypermobility reports somewhat more variation in their responses than the group that does not have extreme hypermobility. The 25^th^ percentile change among those with extreme hypermobility is negative for all four symptoms (suggesting less hypermobility currently than before the got COVID-19), but 0 for those without extreme hypermobility for all four symptoms. The 75^th^ percentile change is positive for all four symptoms among those with extreme hypermobility but only positive for two symptoms among those without extreme hypermobility.

*Supplemental Table 1. Change in Average Hypermobility Symptoms Scores from Before COVID to Currently among Respondents who Report Long COVID Sample, by extreme hypermobility status*

| Descriptives | | | | | | | | | | |
| --- | --- | --- | --- | --- | --- | --- | --- | --- | --- | --- |
|  | | **Extreme Hypermobility** | | **Change in Bendiness/Double-Jointedness** | | **Change in Clicky or Dislocating Joints** | | **Change in Bruising Easily** | | **Change in Unusually Stretchy Skin** |
| N |  | Not extremely hypermobile |  | 309 |  | 309 |  | 309 |  | 309 |
|  |  | Extremely hypermobile |  | 43 |  | 43 |  | 43 |  | 43 |
| Missing |  | Not extremely hypermobile |  | 0 |  | 0 |  | 0 |  | 0 |
|  |  | Extremely hypermobile |  | 0 |  | 0 |  | 0 |  | 0 |
| Mean |  | Not extremely hypermobile |  | -1.13 |  | 2.77 |  | 7.97 |  | 2.14 |
|  |  | Extremely hypermobile |  | -14.3 |  | -5.60 |  | 8.51 |  | 3.44 |
| Standard deviation |  | Not extremely hypermobile |  | 19.5 |  | 16.7 |  | 22.4 |  | 15.4 |
|  |  | Extremely hypermobile |  | 31.4 |  | 34.1 |  | 27.1 |  | 22.2 |
| 25th percentile |  | Not extremely hypermobile |  | 0.00 |  | 0.00 |  | 0.00 |  | 0.00 |
|  |  | Extremely hypermobile |  | -25.5 |  | -16.0 |  | -3.00 |  | -4.50 |
| 50th percentile |  | Not extremely hypermobile |  | 0.00 |  | 0.00 |  | 0.00 |  | 0.00 |
|  |  | Extremely hypermobile |  | -3.00 |  | -2.00 |  | 2.00 |  | 0.00 |
| 75th percentile |  | Not extremely hypermobile |  | 0.00 |  | 1.00 |  | 11.0 |  | 0.00 |
|  |  | Extremely hypermobile |  | 0.50 |  | 8.00 |  | 21.5 |  | 3.00 |
